# Efficacy of face mask in preventing respiratory virus transmission: a systematic review and meta-analysis

**DOI:** 10.1101/2020.04.03.20051649

**Authors:** Mingming Liang, Liang Gao, Ce Cheng, Qin Zhou, John Patrick Uy, Kurt Heiner, Chenyu Sun

**Affiliations:** Department of Epidemiology and Health Statistics, School of Public Health, Anhui Medical University, Hefei 230032, Anhui, P.R. China; Center for Evidence-Based Practice, Anhui Medical University, Hefei 230032, Anhui, P.R. China; Center of Experimental Orthopaedics, Saarland University Medical Center, Kirrberger Straße Building 37-38, D-66421 Homburg/Saar, Germany; Department of Internal Medicine, Cape Fear Valley Medical Center, Fayetteville 28304, NC, USA; Mayo Clinic, Rochester, MN 55905, USA; AMITA Health Saint Joseph Hospital Chicago, Chicago 60657, Illinois, USA; Dignity Health Mercy Hospital, Merced 95340, CA, USA

**Keywords:** Facemask, Respiratory virus, Influenza, SARS-CoV, SARS-CoV-2, Prevention

## Abstract

**Background:** Conflicting recommendations exist related to whether masks have a protective effect on the spread of respiratory viruses.

**Methods:** The Preferred Reporting Items for Systematic Reviews and Meta-Analysis (PRISMA) statement was consulted to report this systematic review. Relevant articles were retrieved from PubMed, Web of Science, ScienceDirect, Cochrane Library, and Chinese National Knowledge Infrastructure (CNKI), VIP (Chinese) database.

**Results:** A total of 21 studies met our inclusion criteria. Meta-analyses suggest that mask use provided a significant protective effect (OR=0.35 and 95% CI=0.24-0.51). Use of masks by healthcare workers (HCWs) and non-healthcare workers (Non-HCWs) can reduce the risk of respiratory virus infection by 80% (OR=0.20, 95% CI=0.11-0.37) and 47% (OR=0.53, 95% CI=0.36-0.79). The protective effect of wearing masks in Asia (OR=0.31) appeared to be higher than that of Western countries (OR=0.45). Masks had a protective effect against influenza viruses (OR=0.55), SARS (OR=0.26), and SARS-CoV-2 (OR=0.04). In the subgroups based on different study designs, protective effects of wearing mask were significant in cluster randomized trials and observational studies.

**Conclusions:** This study adds additional evidence of the enhanced protective value of masks, we stress that the use masks serve as an adjunctive method regarding the COVID-19 outbreak.

## Introduction

Facemasks are recommended for diseases transmitted through droplets and respirators for respiratory aerosols, yet recommendations and terminology vary between guidelines. The concepts of droplet and airborne transmission that are entrenched in clinical practice recently are more complex than previously thought. The concern is now increasing in the face of the Coronavirus Disease 2019 (COVID-19) pandemic [1]. The spread of respiratory viral infections (RVIs) occurs primarily through contact and droplet routes. And new evidence suggests severe acute respiratory syndrome coronavirus 2 (SARS-CoV-2) can remain viable and infectious in aerosols for hours [2]. Therefore, the use of masks as appropriate personal protective equipment (PPE) is often considered when preventing the spread of respiratory infections. Experimental data shows that the micropores of mask block dust particles or pathogens that are larger than the size of micropores. For example, the micropores of N95 masks materials are only 8 microns in diameter, which can effectively prevent the penetration of virions [3–5].

Although the aforementioned studies support the potential beneficial effect of masks, the substantial impact of masks on the spread of laboratory-diagnosed respiratory viruses remains controversial [6]. Smith et al. indicated that there were insufficient data to determine definitively whether N95 masks are superior to surgical masks in protecting healthcare workers (HCWs) against transmissible acute respiratory infections in clinical settings [7]. Another meta-analysis suggested that facemask provides a non-significant protective effect (OR=0.53, 95% CI 0.16-1.71, I^2^=48%) against the 2009 influenza pandemic [8]. Xiao et al. concluded that masks did not support a substantial effect on the transmission of influenza from 7 studies [6]. On the contrary, Jefferson et al. suggested that wearing masks significantly decreased the spread of SARS (OR=0.32; 95% CI 0.25-0.40; I^2^=58.4%) [9]. Up to date, existing evidence on the effectiveness of the use of masks to prevent respiratory viral transmission contradicts each other.

Therefore, we performed a systematic review and meta-analysis to evaluate the effectiveness of the use of masks to prevent laboratory-confirmed respiratory virus transmission.

## Methods

### Identification and selection of studies

The Preferred Reporting Items for Systematic Reviews and Meta-Analysis (PRISMA) statement was consulted to report this systematic review. Regarding this meta-analysis, a comprehensive searching strategy was carefully designed to select eligible studies from multiple electronic databases, including PubMed, Web of Science, Cochrane Library, and Chinese National Knowledge Infrastructure (CNKI), VIP (Chinese) database. All included studies were published before March 2020. Relevant Chinese technical terms for the Chinese databases were used to search for published articles (see **Appendix 1**, for search details). Furthermore, references of all relevant articles and reviews were retrieved to search for additional eligible studies. Articles just providing abstracts were excluded. After deleting duplicates, all abstracts and titles were filtered independently by two reviewers to remove the irrelevant articles. We downloaded and read the full text of the potential research related to the selection criteria to incorporate systematic reviews. Reviewers compared and discussed the results. If a discussion by the two reviewers did not result in an agreement, then the third party was called upon to create consensus.

### Inclusion and exclusion criteria

The studies meeting the following criteria were included: (1) concerning the relationship between the face mask and preventing RVIs; (2) diagnosis of respiratory virus must have laboratory evidence, or the local clinical diagnostic criteria are applied during an acute large-scale infectious disease when laboratory evidence might be not available; (3) providing complete data of cases and controls for calculating an odds ratio (OR) with 95% confidence interval (CI); (4) no language restrictions applied. The exclusive criteria were as follows: (1) insufficient data to ascertain the adjusted ORs; (2) conferences/meetings abstracts, case reports, editorials, and review articles; (3) duplicate publication or overlapping studies.

## Study quality assessment

The Newcastle-Ottawa Scale (NOS) was used to evaluate the quality of the case-control study and cohort study: study ratings of seven to nine stars corresponded to high-quality, five to six stars to moderate quality, and four stars or less to low quality [10]. The Jadad scale was used to evaluate the quality of randomized controlled study: study ratings of three to five corresponded to high-quality, and two or less to low quality [11]. Three members of the review team completed assessments independently. The disagreements were resolved by discussion.

### Statistical analysis

The association of mask use with subsequent RVIs was assessed with odds ratios (OR) with a 95% confidence interval (CI). P values less than 0.05 were considered statistically significant. Considering the potential for between-study heterogeneity, subgroup analyzes were carried out based on stratification by HCWs, countries, virus types, and study designs. Sensitivity analysis was performed by omitting individual studies to assess the stability of the meta-analysis. The heterogeneity was assessed using the I^2^ statistic. The heterogeneity was considered insignificance when P > 0.10 and I^2^ < 50%. If the study lacked heterogeneity, the pooled OR estimate was calculated using the fixed-effects model, otherwise the random-effects model was used [12]. Begg’s and Egger’s test were performed to quantitatively analyze the potential publication bias by Stata (version 14.0; Stata Corp, College Station, TX) software. The P values of Begg’s and Egger’s test more than 0.05 implied no obvious publication bias in this meta-analysis [13, 14]. The meta-analysis was performed using Revman 5.3.5 (http://tech.cochrane.org/revman) [15].

## Results

### Characteristics of eligible studies

A flow diagram of the literature search and related screening process is shown in Figure 1. A total of 21 studies met our inclusion criteria [4, 16–35], including 13 case-control studies, 6 cluster randomized trials, 2 cohort study (Table 1). Among them, 11 studies were conducted in China (includes 4 studies from Hong Kong, China), 6 in Western countries, and 4 in other Asian countries. 4 studies investigated patients with respiratory virus, 7 studies investigated Severe acute respiratory syndrome coronavirus (SARS-CoV), 12 studies investigated influenza virus including 5 investigating the H1N1 virus, and 1 study investigated SARS-CoV-2. All patients had laboratory evidence or met local clinical diagnostic criteria during an acute large-scale infectious disease crisis. There were 12 studies targeting HCWs, the remaining 8 studies investigated non-healthcare professional populations.

**Table 1.**
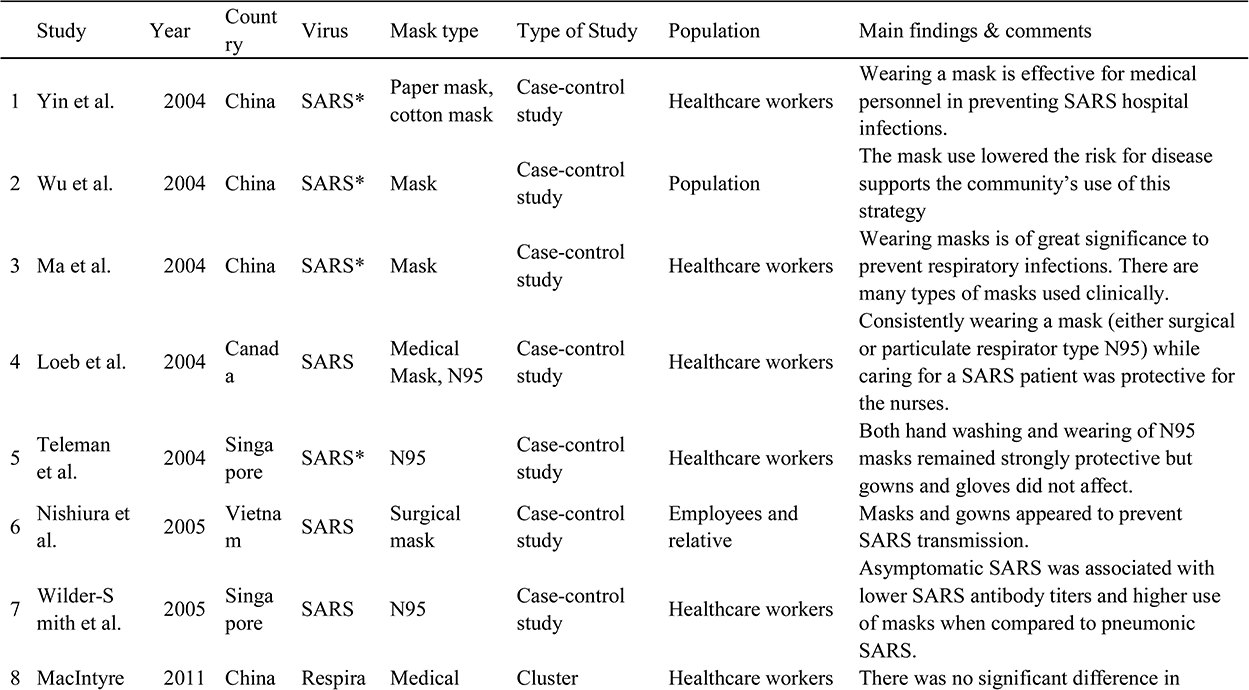
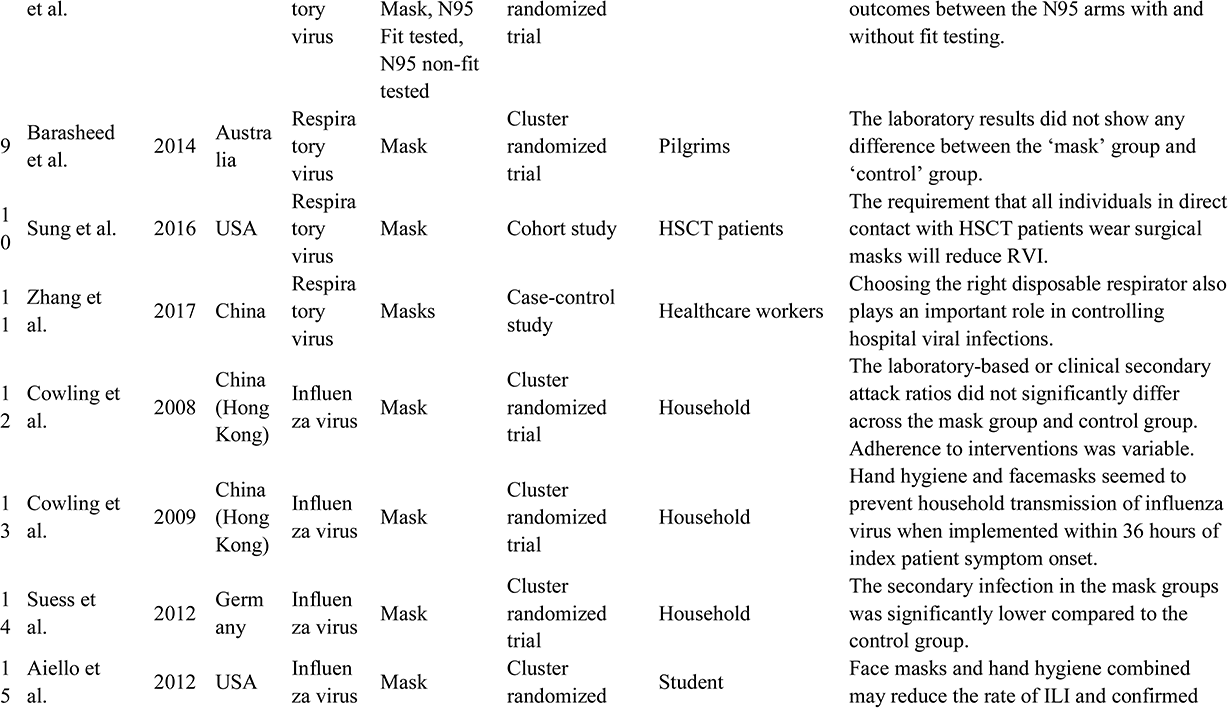
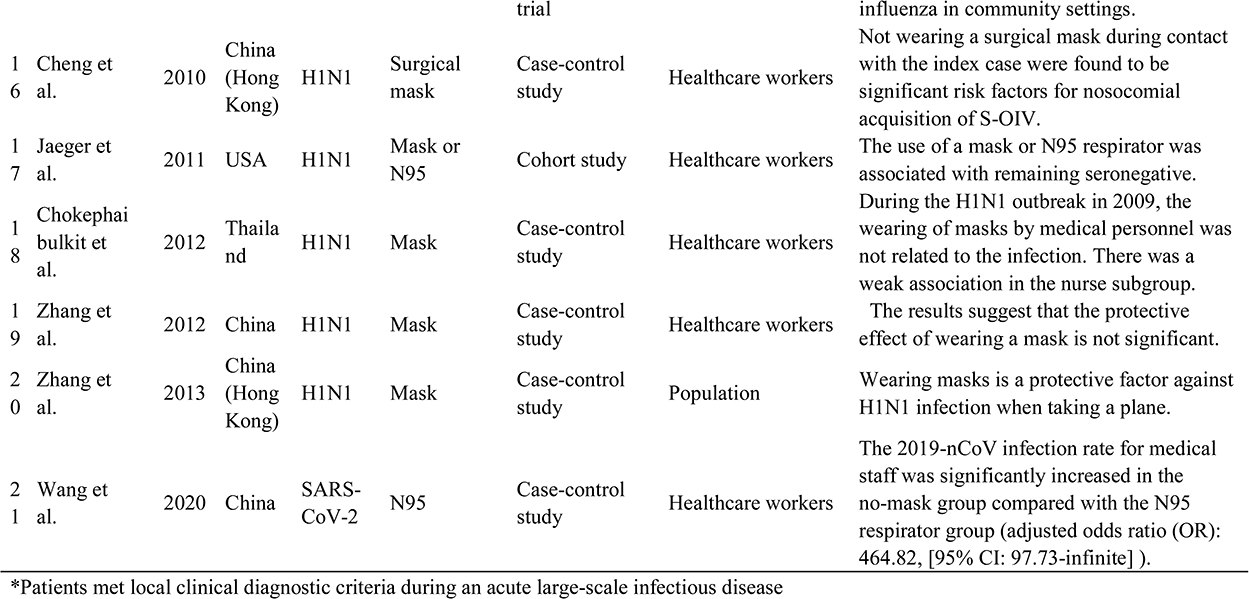
Characteristics of eligible studies

| Study | Year | Country | Virus | Mask type | Type of Study | Population | Main findings & comments |
| --- | --- | --- | --- | --- | --- | --- | --- |
| 1 Yin et al. | 2004 | China | SARS* | Paper mask, cotton mask | Case-control study | Healthcare workers | Wearing a mask is effective for medical personnel in preventing SARS hospital infections. |
| 2 Wu et al. | 2004 | China | SARS* | Mask | Case-control study | Population | The mask use lowered the risk for disease supports the community's use of this strategy |
| 3 Ma et al. | 2004 | China | SARS* | Mask | Case-control study | Healthcare workers | Wearing masks is of great significance to prevent respiratory infections. There are many types of masks used clinically. |
| 4 Loeb et al. | 2004 | Canada | SARS | Medical Mask, N95 | Case-control study | Healthcare workers | Consistently wearing a mask (either surgical or particulate respirator type N95) while caring for a SARS patient was protective for the nurses. |
| 5 Teleman et al. | 2004 | Singapore | SARS* | N95 | Case-control study | Healthcare workers | Both hand washing and wearing of N95 masks remained strongly protective but gowns and gloves did not affect. |
| 6 Nishiura et al. | 2005 | Vietnam | SARS | Surgical mask | Case-control study | Employees and relative | Masks and gowns appeared to prevent SARS transmission. |
| 7 Wilder-Smith et al. | 2005 | Singapore | SARS | N95 | Case-control study | Healthcare workers | Asymptomatic SARS was associated with lower SARS antibody titers and higher use of masks when compared to pneumonic SARS. |
| 8 MacIntyre | 2011 | China | Respira | Medical | Cluster | Healthcare workers | There was no significant difference in |

|  |  |  |  |  |  |  |  |  |
| --- | --- | --- | --- | --- | --- | --- | --- | --- |
|  | et al. |  |  | tory virus | Mask, N95<br>Fit tested,<br>N95 non-fit tested | randomized trial |  | outcomes between the N95 arms with and without fit testing. |
| 9 | Barasheed et al. | 2014 | Australia | Respiratory virus | Mask | Cluster randomized trial | Pilgrims | The laboratory results did not show any difference between the 'mask' group and 'control' group. |
| 10 | Sung et al. | 2016 | USA | Respiratory virus | Mask | Cohort study | HSCT patients | The requirement that all individuals in direct contact with HSCT patients wear surgical masks will reduce RVI. |
| 11 | Zhang et al. | 2017 | China | Respiratory virus | Masks | Case-control study | Healthcare workers | Choosing the right disposable respirator also plays an important role in controlling hospital viral infections. |
| 12 | Cowling et al. | 2008 | China (Hong Kong) | Influenza virus | Mask | Cluster randomized trial | Household | The laboratory-based or clinical secondary attack ratios did not significantly differ across the mask group and control group. Adherence to interventions was variable. |
| 13 | Cowling et al. | 2009 | China (Hong Kong) | Influenza virus | Mask | Cluster randomized trial | Household | Hand hygiene and facemasks seemed to prevent household transmission of influenza virus when implemented within 36 hours of index patient symptom onset. |
| 14 | Suess et al. | 2012 | Germany | Influenza virus | Mask | Cluster randomized trial | Household | The secondary infection in the mask groups was significantly lower compared to the control group. |
| 15 | Aiello et al. | 2012 | USA | Influenza virus | Mask | Cluster randomized trial | Student | Face masks and hand hygiene combined may reduce the rate of ILI and confirmed influenza in community settings. |
| 1<br>6 | Cheng et al. | 2010 | China (Hong Kong) | H1N1 | Surgical mask | Case-control study | Healthcare workers | Not wearing a surgical mask during contact with the index case were found to be significant risk factors for nosocomial acquisition of S-OIV. |
| 1<br>7 | Jaeger et al. | 2011 | USA | H1N1 | Mask or N95 | Cohort study | Healthcare workers | The use of a mask or N95 respirator was associated with remaining seronegative. |
| 1<br>8 | Chokephai bulkit et al. | 2012 | Thailand | H1N1 | Mask | Case-control study | Healthcare workers | During the H1N1 outbreak in 2009, the wearing of masks by medical personnel was not related to the infection. There was a weak association in the nurse subgroup. |
| 1<br>9 | Zhang et al. | 2012 | China | H1N1 | Mask | Case-control study | Healthcare workers | The results suggest that the protective effect of wearing a mask is not significant. |
| 2<br>0 | Zhang et al. | 2013 | China (Hong Kong) | H1N1 | Mask | Case-control study | Population | Wearing masks is a protective factor against H1N1 infection when taking a plane. |
| 2<br>1 | Wang et al. | 2020 | China | SARS-CoV-2 | N95 | Case-control study | Healthcare workers | The 2019-nCoV infection rate for medical staff was significantly increased in the no-mask group compared with the N95 respirator group (adjusted odds ratio (OR): 464.82, [95% CI: 97.73-infinite] ). |

\*Patients met local clinical diagnostic criteria during an acute large-scale infectious disease

### Quality of studies

Inter-rater agreement of the quality of included studies was strong. Table 2 and 3 summarize the quality evaluations of the included studies. Funnel plots assessing the risk of publication bias are included in figure 2. Neither Begg’s test (z=0.45, p=0.651) nor Egger’s test (t=-0.65, p=0.524) manifested any distinct evidence of the publication bias. The sensitivity analyses did not substantially alter the pooled ORs by excluding one-by-one study, indicating that the meta-analysis was generally robust.

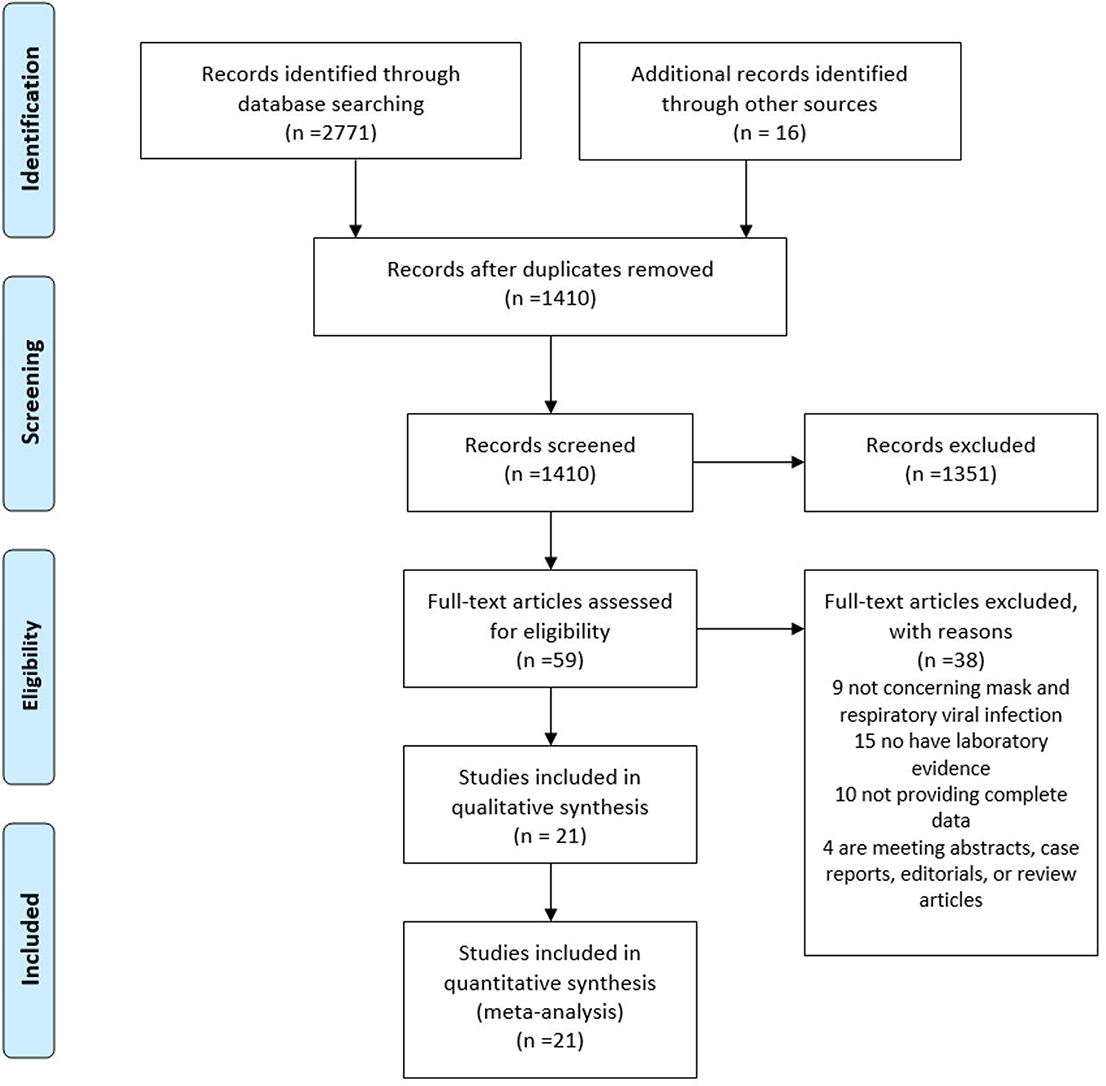

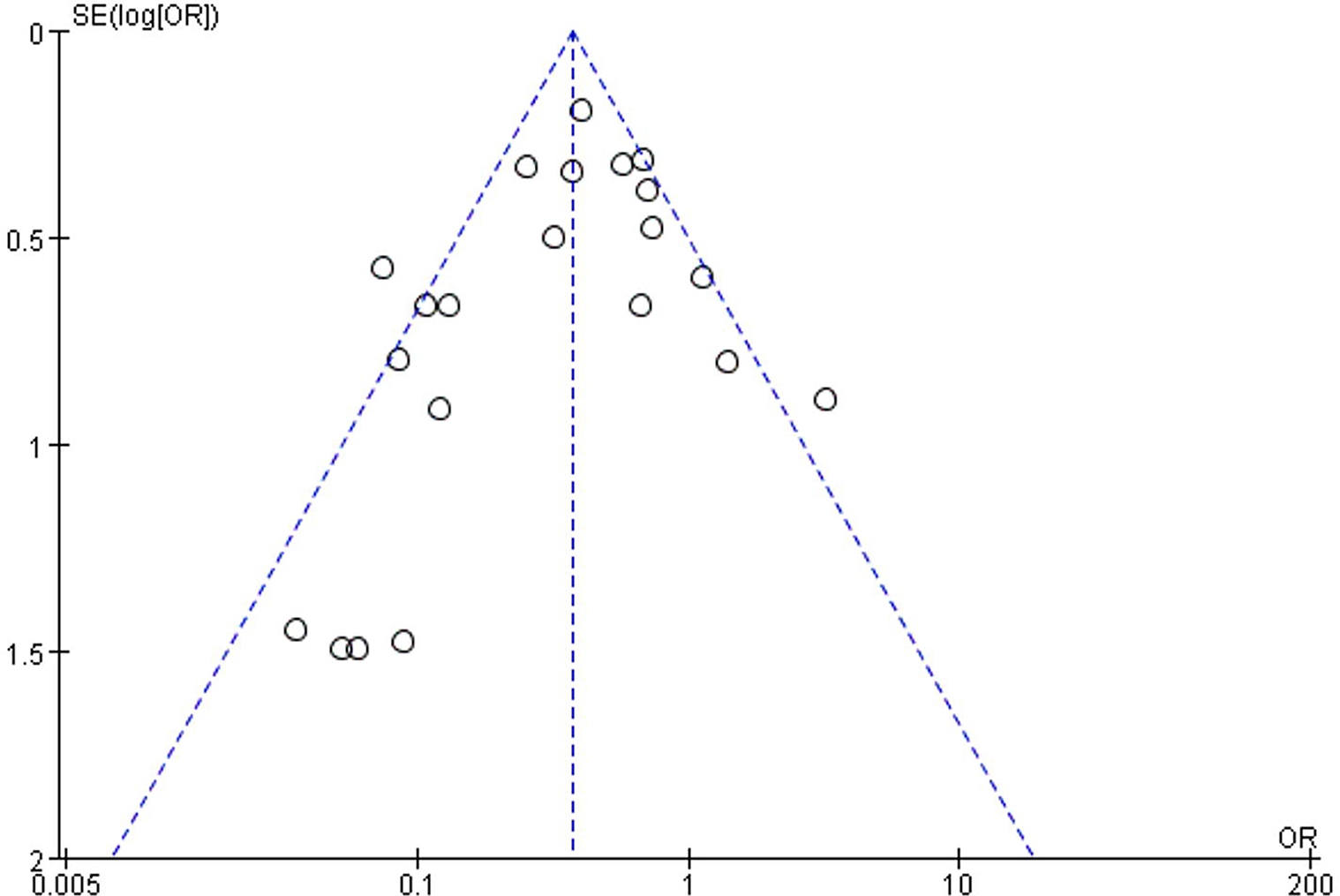

**Table 2.** The quality of the case-control studies and cohort studies

|  | Study | Year | Selection<br>n | Comparability | Outcome | Stars* |
| --- | --- | --- | --- | --- | --- | --- |
| 1 | Yin et al. | 2004 | 3 | 2 | 2 | 7 |
| 2 | Wu et al. | 2004 | 4 | 2 | 2 | 8 |
| 3 | Ma et al. | 2004 | 3 | 2 | 2 | 8 |
| 4 | Loeb et al. | 2004 | 3 | 2 | 2 | 7 |
| 5 | Teleman et al. | 2004 | 3 | 2 | 3 | 8 |
| 6 | Wilder-Smith et al. | 2005 | 3 | 2 | 3 | 8 |
| 7 | Nishiura et al. | 2005 | 4 | 2 | 1 | 7 |
| 8 | Cheng et al. | 2010 | 3 | 2 | 3 | 8 |
| 9 | Jaeger et al. | 2011 | 3 | 2 | 2 | 7 |
| 10 | Chokephaibulkit et al. | 2012 | 3 | 2 | 2 | 7 |
| 11 | Zhang et al. | 2012 | 3 | 2 | 3 | 8 |
| 12 | Zhang et al. | 2013 | 4 | 2 | 1 | 7 |
| 13 | Sung et al. | 2016 | 3 | 2 | 2 | 7 |
| 14 | Zhang et al. | 2017 | 3 | 2 | 1 | 6 |
| 15 | Wang et al. | 2020 | 3 | 1 | 1 | 5 |

\* Scoring by Newcastle-Ottawa Scale

**Table 3.**
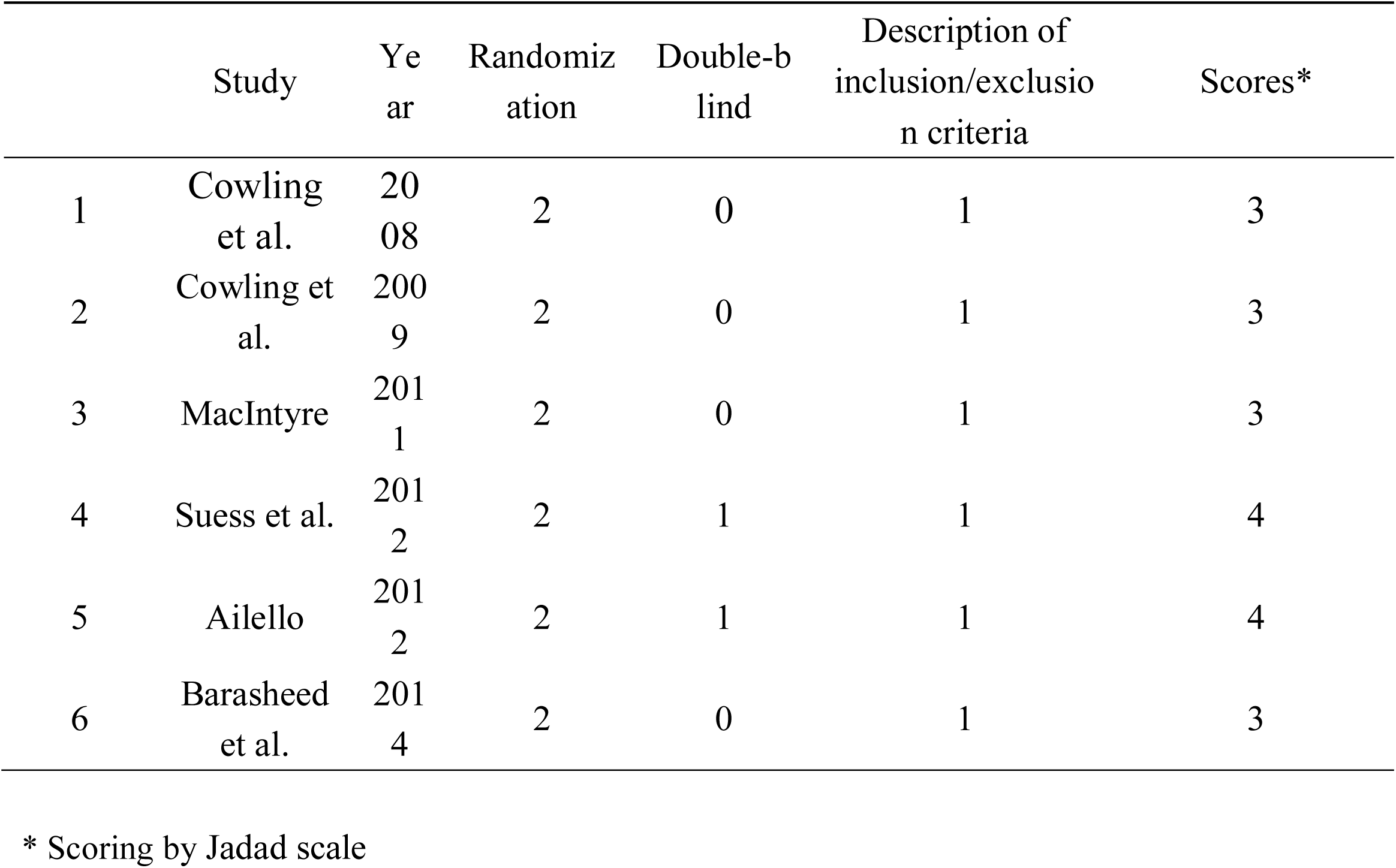
The quality of randomized controlled studies

### Wearing masks reduces the risk of RVIs in general

The 21 studies reporting on the effectiveness of wearing masks included 8,686 participants. In general, masks are effective in preventing the spread of respiratory viruses. After wearing a mask, the risk of contracting RVIs was significantly reduced, with the pooled OR was 0.35 and 95% CI=0.24-0.51 (I^2^=60%, M-H Random-effect model) (Figure 3).

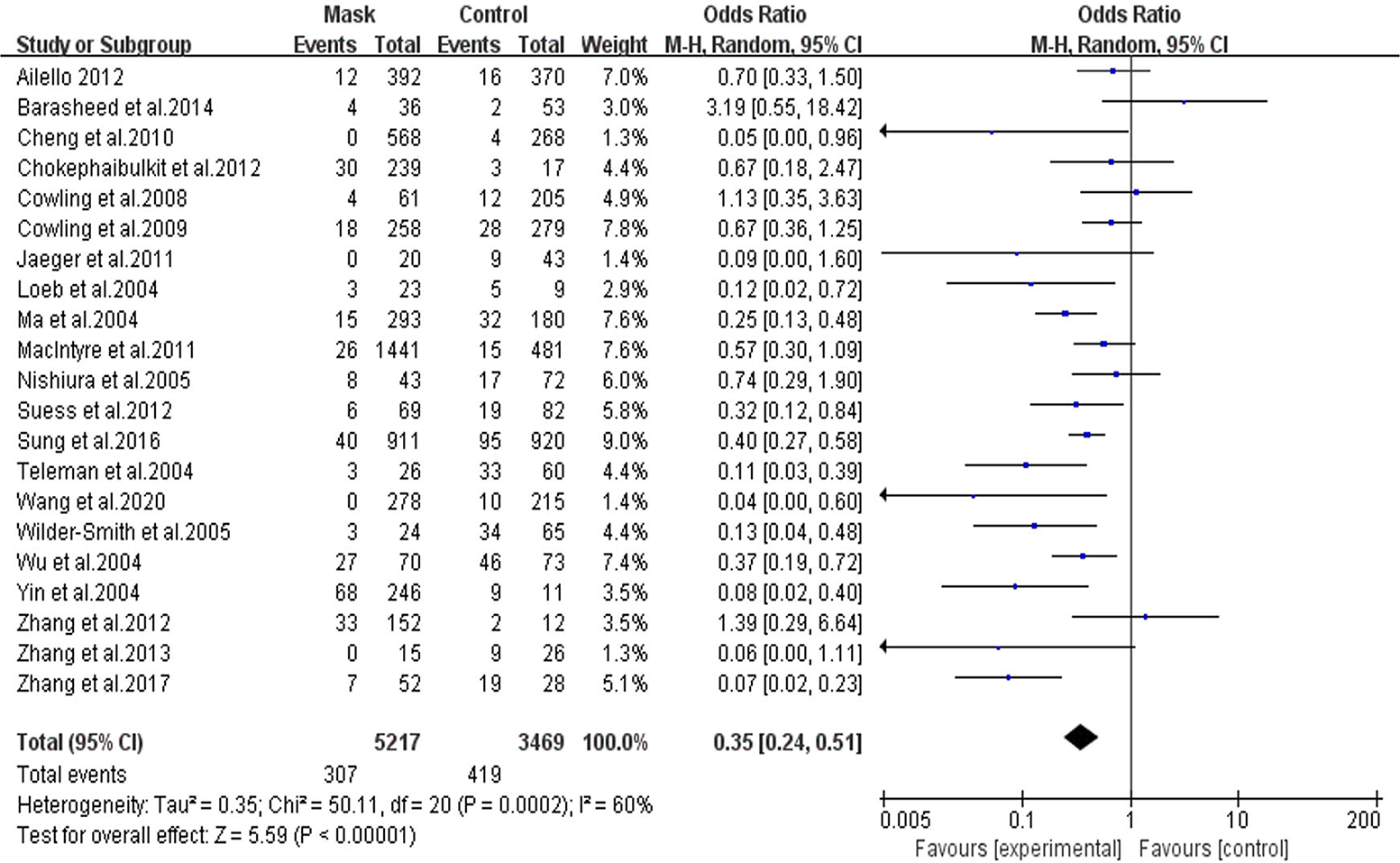

### HCWs vs. non-HCWs

In the subgroup of HCWs only, the protective effect was more obvious, with the pooled OR of 0.20 (95% CI=0.11-0.37, I^2^=59%) (Figure 4). In one study investigating COVID-19, the OR was 0.04 (95%CI=0.00-0.60) [35]. In the subgroup of non-HCW, a protective effect was found with the pooled OR of 0.53 (95% CI=0.36 - 0.79, I^2^=45%). A more detailed analysis found significant effects in both the household subgroup (OR=0.60, 95% CI=0.37-0.97, I^2^=31%), and the non-household subgroup (OR=0.44, 95% CI=0.33-0.59, I^2^=54%) (Figure 5). 1 study included both health care workers and family members of patients, with the OR of 0.74 (95% CI: 0.29-1.90) [22].

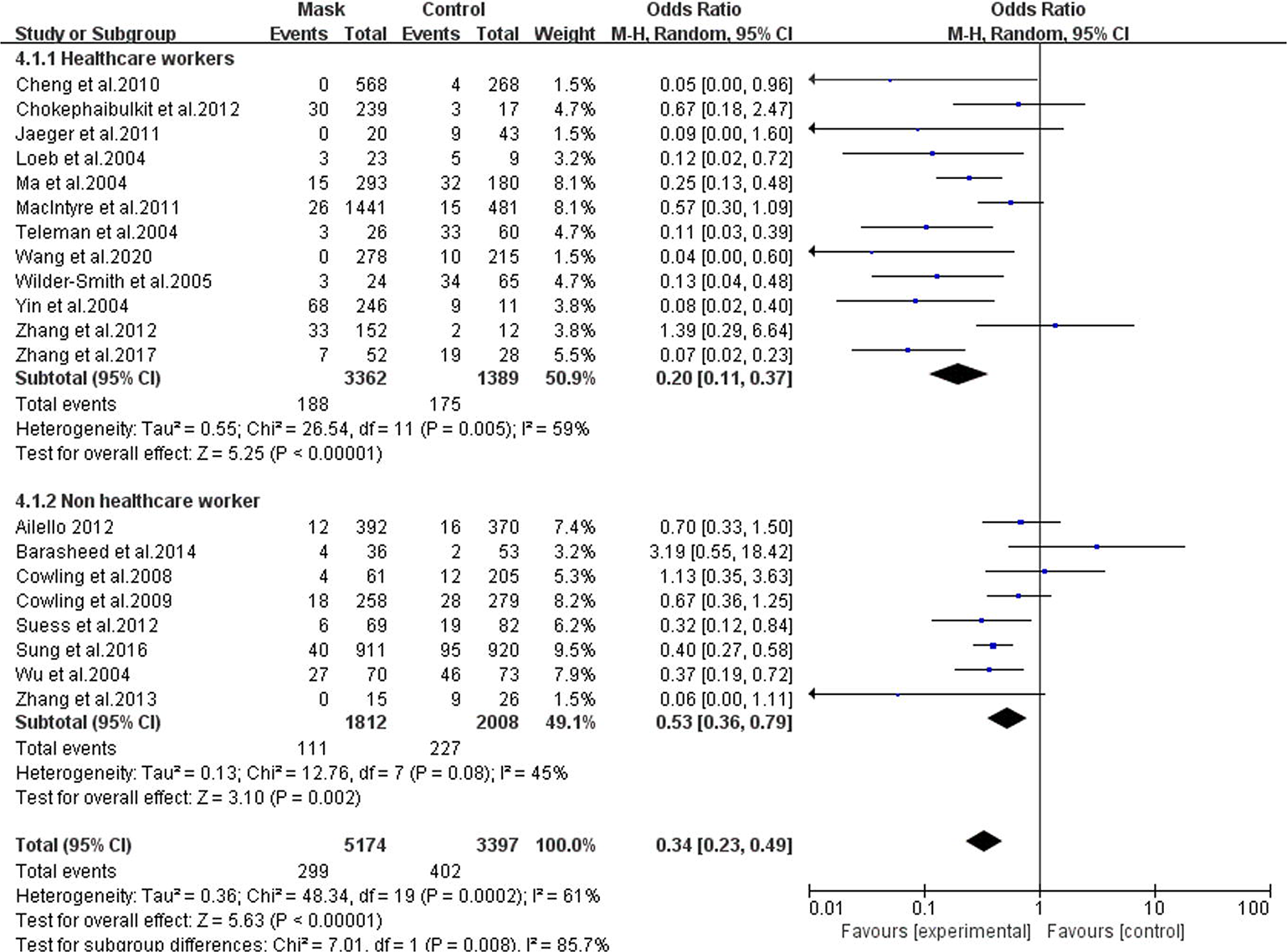

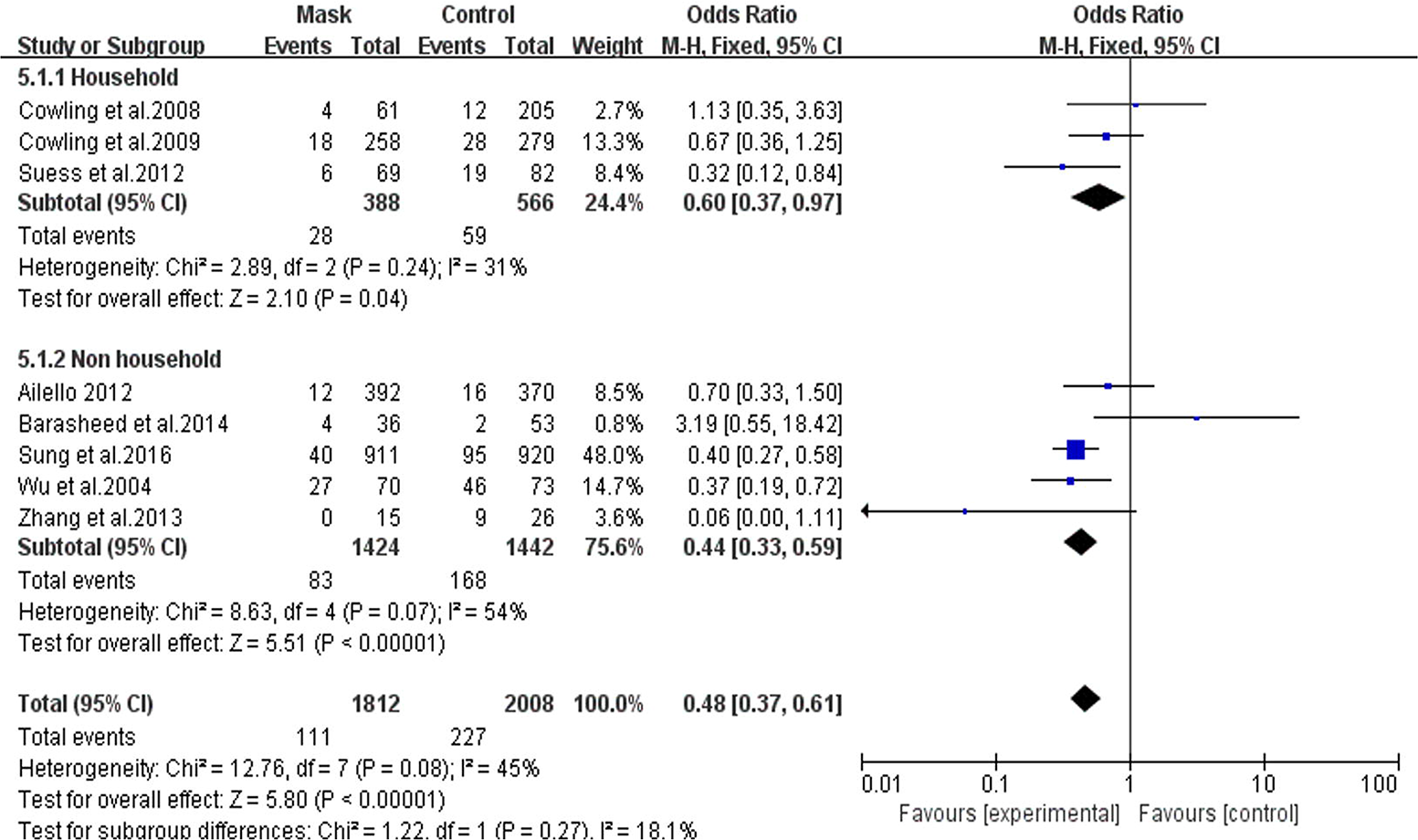

### Subgroup analyses based on areas

By geographic locations, beneficial protective effects of wearing masks were found in Asia (OR=0.31, 95% CI=0.19-0.50, I^2=^65%), and in Western countries (OR=0.45, 95% CI=0.24-0.83, I^2=^51%) (Figure 6). HCWs in both Asia (OR=0.21, 95% CI=0.11-0.41, I^2^=64%) and Western countries (OR=0.11, 95% CI=0.02-0.51, I^2^=0%) can significantly reduce the risk of RVIs by wearing masks (Figure 7). In the non-HCW subgroup, protective effects were found in Western countries (OR=0.46, 95% CI=0.34-0.63, I^2=^57%) and Asia (OR=0.51, 95% CI=0.34-0.78, I^2^=45%) (Figure 8).

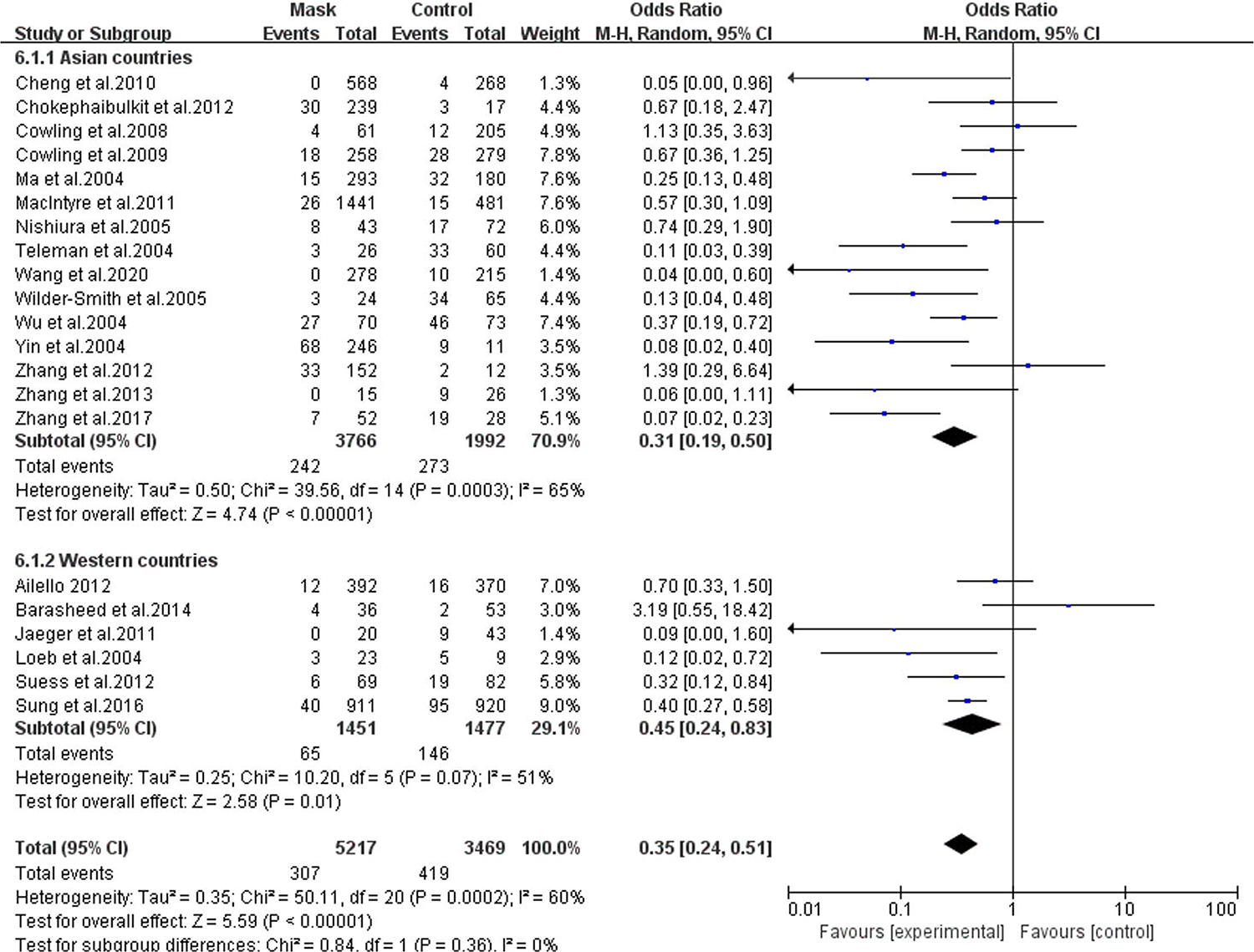

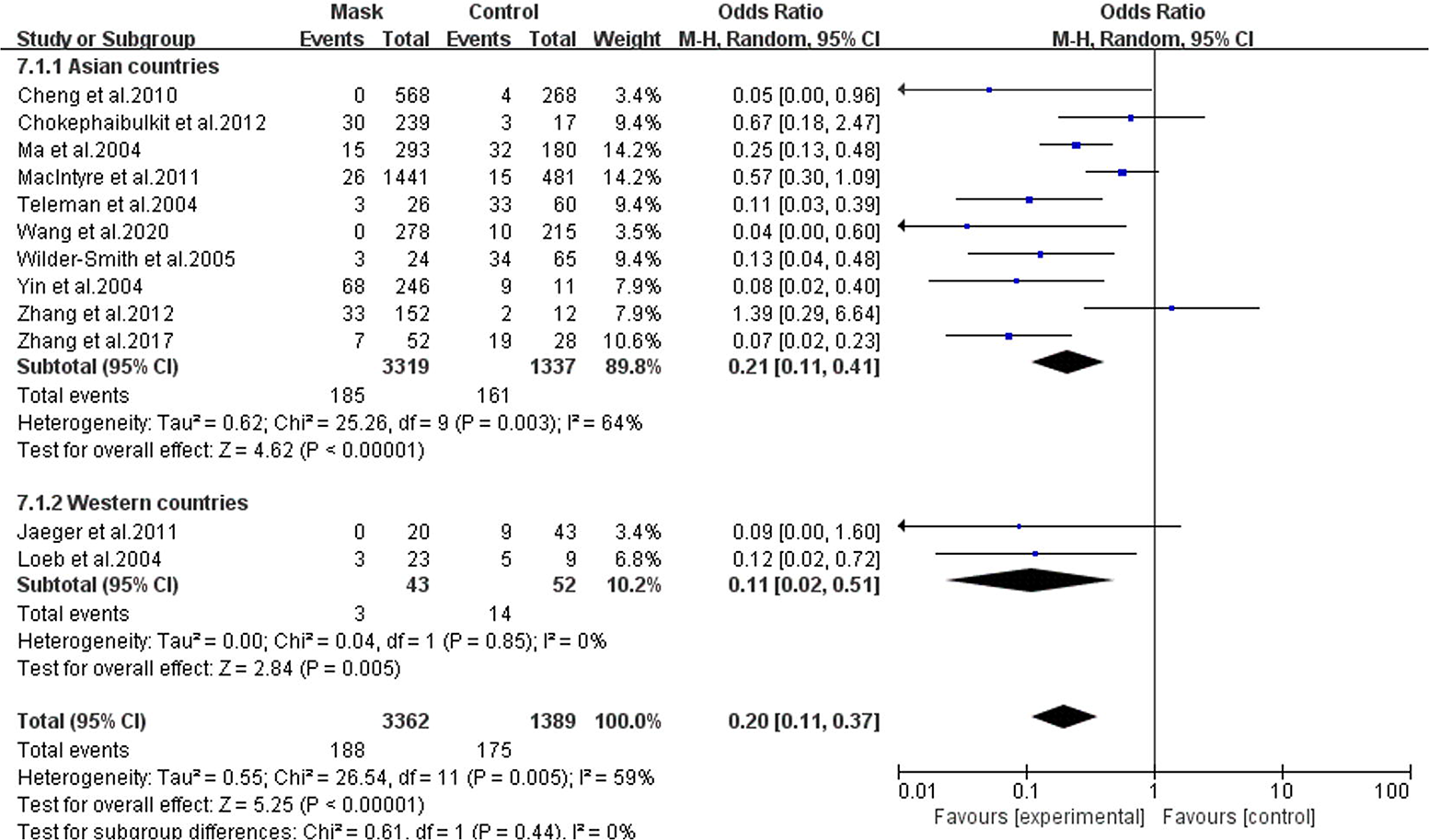

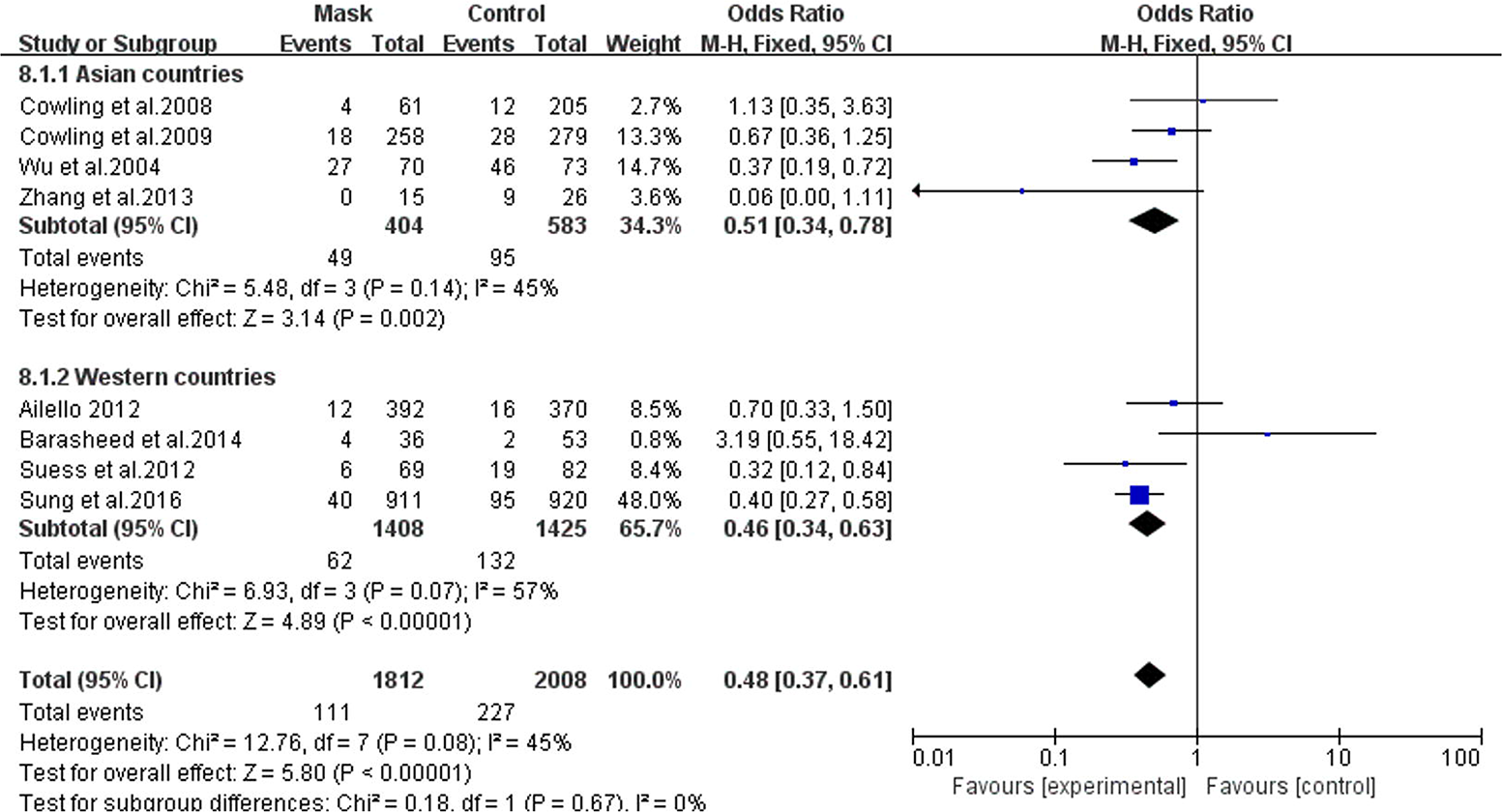

### Subgroup analyses based on different virus types

Masks had a protective effect against influenza viruses (OR=0.55, 95% CI=0.39-0.76, I^2=^27%), SARS (OR=0.26, 95% CI=0.18-0.37, I^2=^47%), and SARS-CoV-2 (OR=0.04, 95% CI=0.00-0.60, I^2=^0%) (Figure 9). However, no significant protective effects against H1N1 was shown (OR=0.30, 95% CI=0.08-1.16, I^2=^51%) (Figure 10).

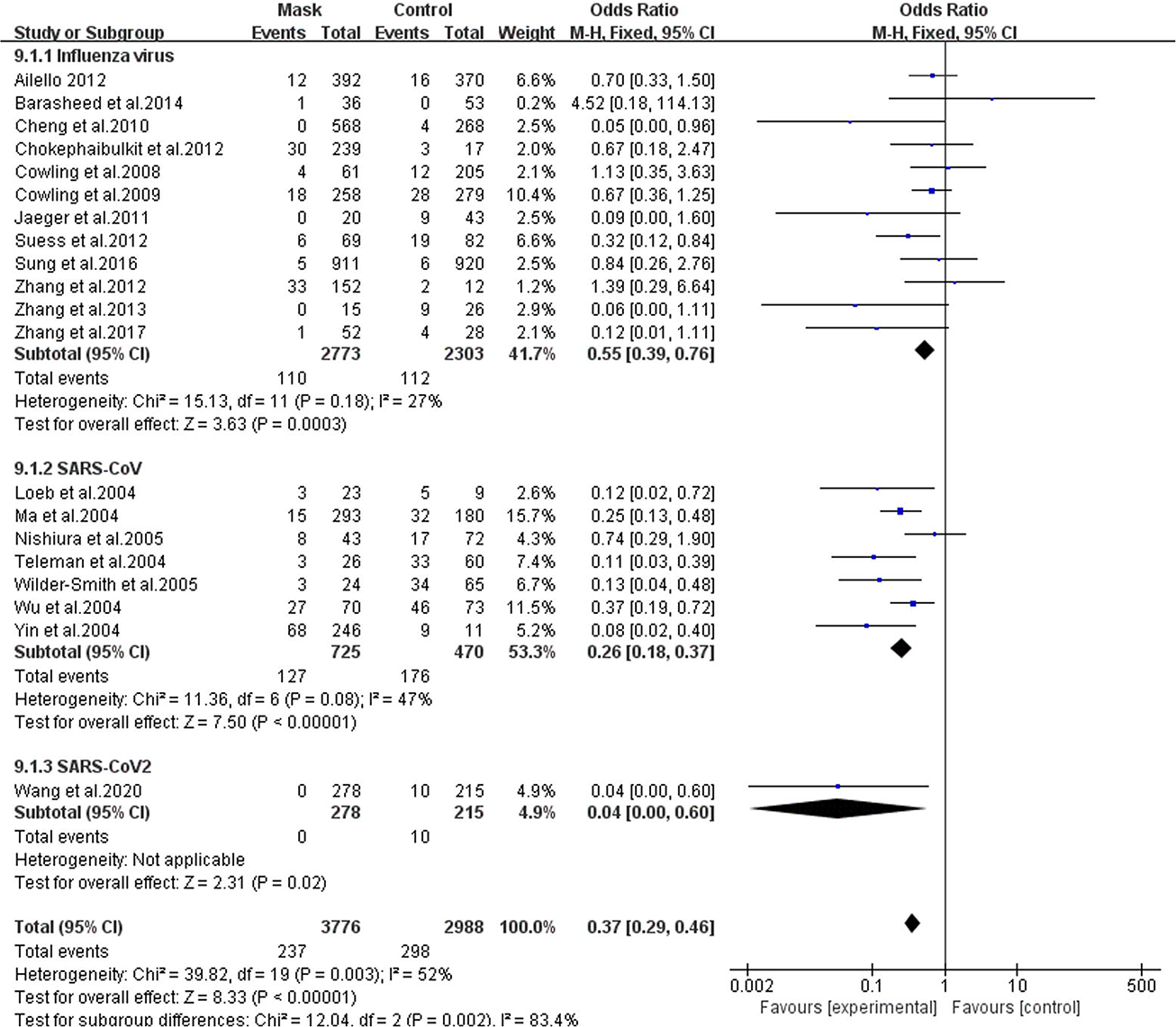

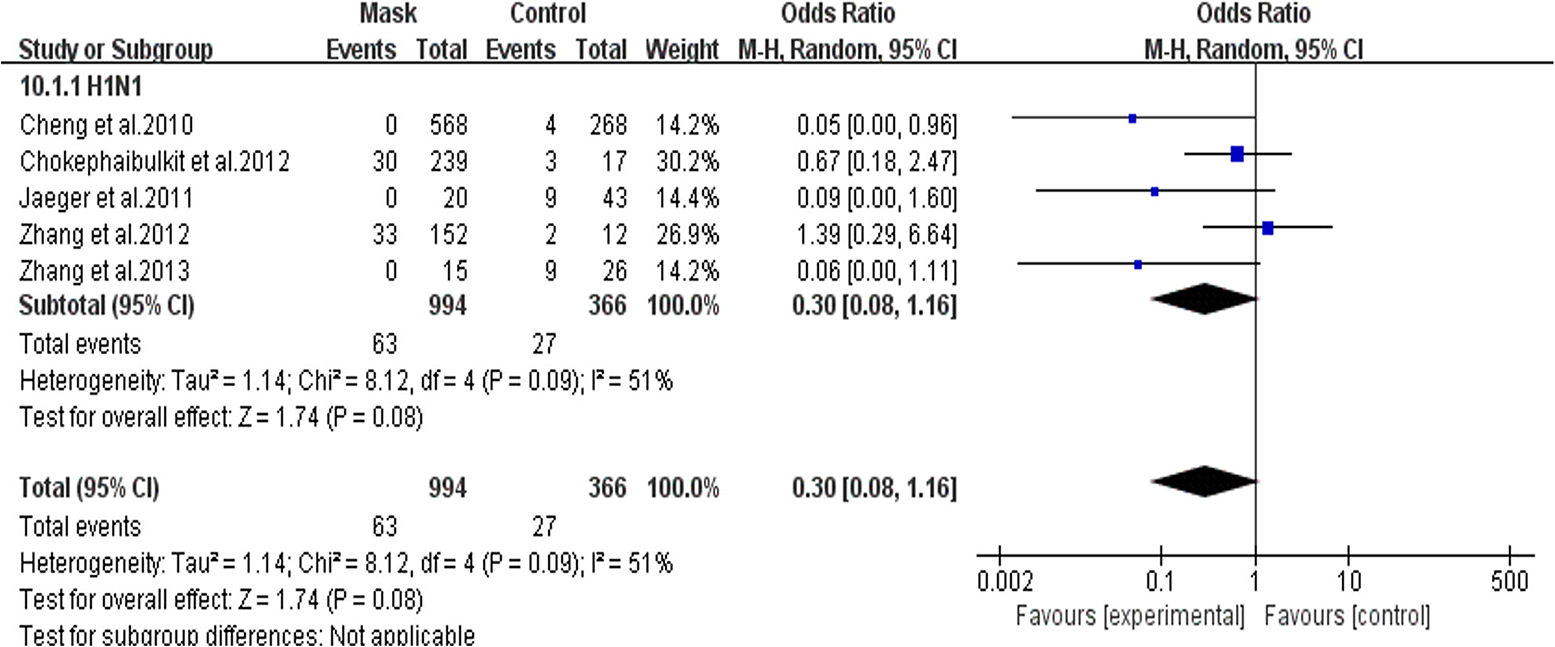

### Subgroup analyses based on different study designs

In the subgroups based on different study designs, protective effects of wearing mask were significant in cluster randomized trials (OR=0.67, 95% CI=0.45-0.98, I^2^=20%) and observational studies (OR=0.24, 95% CI=0.15-0.38, I^2=^54%) (Figure 11).

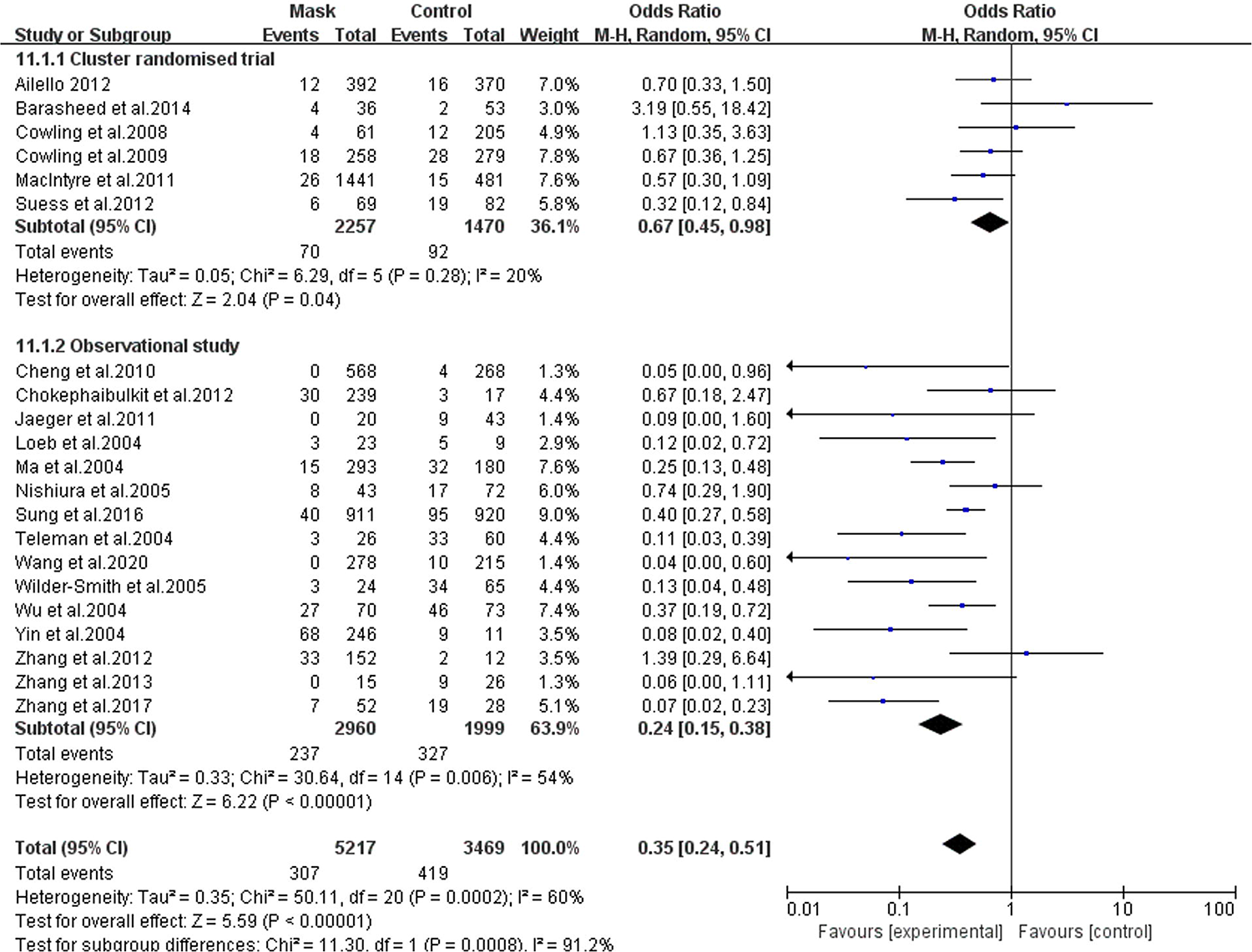

## Discussion

This meta-analysis of all 21 available articles provides the most current evidence to date on the efficacy of masks in preventing the transmission of RVIs. The physical characteristics of the mask can prevent the respiratory tract from contacting the outside virus, thereby reducing the risk of infection with respiratory diseases. The latest research by Leung et al. found that surgical masks significantly reduced the detection of influenza virus RNA in respiratory droplets and coronavirus RNA in aerosols, and there was a tendency to reduce coronavirus RNA in respiratory droplets [36]. Our study shows that masks worn by non-infected people can effectively prevent the spread of respiratory viruses and reduce the overall risk of respiratory virus infection by 65%. This result is similar to Jefferson’s meta-analysis in 2010, which suggested that wearing masks were highly effective in preventing the spread of SARS (OR=0.32, 95%CI:0.25 −0.40) [9]. This is especially instructive during periods of high-risk RVIs, such as the current COVID-19 pandemic [1].

### Wearing mask has protective effects for both HCWs and non-HCWs

During current COVID-19 pandemic [1], it is pivotal to protect the HCWs from getting infected. HCWs are expected to work long hours with often inadequate resources and under significant pressure, facing the dangers inherent in close contact with index-patients during outbreak [37]. Through February 21, 2020, a total of 1,716 HCWs in China have become infected by SARS-CoV-2 and 5 have died [38]. In Italy, more than 2,600 healthcare workers have been infected by March 19, 2020, accounting for 8.3% of the country’s total cases [39]. According to our analysis, wearing masks significantly reduced the risk of infection among HCWs by 80%. It is noteworthy that, none of the 278 HCWs wearing N95 masks in quarantined areas were infected by SARS-CoV-2 yet, 10 of the 215 HCWs who did not wear masks in the open areas were infected [35]. Therefore, wearing proper PPE including N95 masks at all clinical settings are likely to provide great benefits for HCWs during current COVID-19 pandemic.

The subgroup analysis found that it is effective not only for HCWs who are in close contact with patients but also for the non-HCWs. In non-household settings, wearing masks reduced the risk by 55%. Moreover, significant protective effects were found in the study conducted in the general population [17], indicating the potential benefits of wearing masks for the general public. Although laboratory-confirmed virus results show no difference between the mask group and the control group in a study investigating the wearing of masks by pilgrims, wearing masks reduced the risk of influenza-like illness when people gather [4]. This difference between laboratory-confirmed cases and clinically diagnosed influenza-like illness cases were likely due to an underdiagnosis of real cases caused by too few nasal swabs collected for laboratory confirmation. Zhang et al. conducted a case-control study and found that none of the passengers always wearing masks on the international flight were infected with H1N1 [32], further suggesting that wearing masks properly may be protective when using public transport [40]. More importantly, protective benefits were also reported in hematopoietic stem cell transplant (HSCT) patients [33]. Besides, Sokol’s study also found a reduced risk of hospital-acquired RVIs by putting surgical masks on all workers and visitors in every patient room on the bone marrow transplant unit [41]. People with older age, immunosuppressed state and underlying comorbidities such as hypertension, diabetes, cardiovascular disease, lung disease, and malignancy are more prone to become severely ill when infected by SARS-CoV-2 [42–44]. Therefore, providing appropriate safety protection measures for them, such as wearing masks, may reduce the risk of infection during current COVID-19 outbreak. In addition, those who have close contact with the elderlies, immunosuppressive patients, and people with underlying comorbidities should consider wearing masks as well.

Protective effects were also found among household setting showing a 40% reduced risk of RVIs. However, one household study included in our analysis found that during the follow-up period, only 21% of face mask arm family contacts often or always wear masks, and it failed to show protective effects of wearing masks [23]. In another study conducted by Cowling et al. [24], low facemask adherence among household contacts was also reported, which might explain the poor protective effects. On the contrary, Suess et al. reported a good compliance, which showed a significant protective effect [29]. These findings implicated that proper use of masks has an impact on the effectiveness of preventing RVIs. Given that most people in household settings were unlikely to strictly follow hand hygiene and mask use recommendations [23], it is more critical to re-evaluate the strategy of self-quarantine or self-isolation at home during current COVID-19 outbreak due to higher risk of family cluster infection [45, 46].

### Wearing mask has protective effects against influenza, SARS, and COVID-19

The risk of influenza, SARS, and COVID-19 infection reduced by 45%, 74%, and 96%, respectively. This is similar to the result of Jefferson’s meta-analysis [9]: masks have a significant protective effect on SARS infection. However, Xiao’s results concluded that the protection of masks against the influenza virus was not significant [6]. Nevertheless, one study included by Xiao et al. was complicated by the arrival of the 2009 H1N1 influenza pandemic and the subsequent national hygiene campaign that prompted behavioral changes in the control group, making it difficult to obtain a convincing result [47].

Also, our results of sub-group analysis showed an insignificant reduction of risk of H1N1 by wearing masks, which could be explained by the limitations of the included studies including relatively small sample size, and confounding factors such as prior influenza vaccinations. Jeager et al. 2009 indicated that overall PPE use among HCWs was low as more than 25% reported never wearing PPE and only 17% reported wearing masks with every H1N1 patient encounter, which could significantly lower the sample size of data being analyzed [27]. Also, the same study indicated that majority of HCWs had received seasonal influenza vaccination, which could play a role of confounding factor contributing to protective effects toward control group. Additionally, it was noted that during acute outbreak of H1N1, specific prevention recommendations and measures lagged behind H1N1 exposures. This could suggest that HCWs may already have been infected before wearing masks, further decreasing the powers of data collected. Regarding the relationship of different medical fields and practice settings and influenza, Santo el al mentioned that physicians and registered nurses had higher risks of infection compared with outpatient and allied health staff, which could be the result of a higher risk of exposures [48]. However, Jeagers did not conclude the same findings, which could be explained by poor techniques of using PPE (such as poorly fitted N95 masks) among allied health staff [27].

### Wearing mask has protective effects in both Asian countries and Western Countries

Due to current controversial guidelines between different countries and areas, regarding the general public wearing masks. We also analyzed its effects based on different geographic locations, showing that wearing masks does provide protective effects in both Asian countries and western countries by 69% and 55%, respectively. Among HCWs, it reduced the risk in both Asian and western countries. Among non-healthcare populations, reduced risk of 54% was found in western countries, and a reduced risk of 49% was found in Asia [29]. The demonstrated reduction in risk was not insignificant and would suggest that the proper use of masks might play a significant role in public health efforts to suppress the spread of RVIs, especially during an outbreak.

### Further improvements for original studies are needed in the future

Important knowledge gaps persist. At present, current evidence on the protective efficiency of masks among the general population is still insufficient. Only one study included in our meta-analysis investigated whether people with certain underlying conditions require masks or not [33]. Recall bias in case-control studies seems inevitable [22]. Therefore, high-quality and well-designed RCTs will be desired to investigate the actual protective effectiveness of masks. Although RCT, in general, is the best study design for assessing the effectiveness of interventions [11], it should be noted that many cluster randomized trials are significantly different from ordinary RCTs (control, randomization and blind). In addition, cluster RCTs included in our study generally have insufficient trial design and low adherence and compliance to interventions

Further interventional experiments should also pay attention to the delay between the onset of symptoms and the application of interventions, otherwise, the efficacy of non-drug interventions may be underestimated, or the statistical capacity to detect significant differences may be lacking. Research on enclosed spaces, such as transportation, is relatively rare [32]. Droplet-borne and airborne viruses are likely to cause large-scale infections among passengers sharing closed transportation [49]. In most studies, the detection criteria for virus types are not detailed and clear. Failure to analyze sufficient types of viruses may lead to biased results. For most studies conducted in healthcare settings, there is a lack of data on control subjects without masks. Because it would be unethical to randomly assign health care professionals to non-mask control groups. In this situation, MacIntyre’s method could be adopted: take a convenient selection method, and choose from the hospital’s HCWs in the department without the need for masks as a control [26].

### Strengths and limitations of this study

This study demonstrates the protective effects of masks on HCWs and other populations. The quality of the included studies was relatively high. There are strict inclusion and exclusion criteria, and all patients have laboratory evidence or met local clinical diagnostic criteria during the outbreak. No significant publication bias was found. And detailed analysis in different settings, populations, and areas were conducted to better clarify the effectiveness of wearing masks. At present, the epidemic of COVID-19 has caused widespread concern globally. There is already evidence that SARS-CoV-2 can be transmitted from asymptomatic individuals, which undoubtedly puts great pressure on the protection of non-HCWs [50]. Therefore, it is necessary to summarize the evidence and select effective PPE in the view properly guiding healthcare professionals and general public regarding current COVID-19 pandemic.

This investigation also had several limitations. First, there was a lack of adequately designed and high-quality prospective studies. For clear research purposes, well-planned prospective studies could help us draw stronger evidence to improve understanding of the effectiveness of masks. Second, this article included some studies of SARS patients diagnosed according to clinical diagnostic criteria for SARS due to a low detection rate of RT-PCR [51]. The lack of sufficient virologic evidence may affect our conclusions. However, this effect might not be significant, as 92% of patients with clinical SARS for whom paired sera were available had a >4-fold rise in antibody titer to SARS-CoV [52]. Third, very few studies included in our analysis investigated the effectiveness of wearing masks by the general public, especially during an outbreak, and only one study investigated whether people with one underlying condition require masks or not [33]. Fourth, published studies have shown that different specifications of masks and different wearing methods may affect the protective effect of masks [17, 32]. And when the included studies divided the time/frequency of wearing masks, we only included the group of masks with the longest wearing/highest wearing frequency. This might also ignore effects of the short/infrequent mask-wearing. In addition, the studies we included were mainly conducted in Asia, especially China, and more evidence from other countries is needed to support our views. Last but not least, information about other confounding biases, such as vaccination, hand hygiene, age, gender, and culture, may affect the protective effect of masks.

## Conclusion

Our study showed the effectiveness of wearing masks in protecting HCWs from RVIs, including SARS-CoV-2. Protective effects were also found among non-HCWs in both Western and Asian countries. However, more evidence is still needed to better clarify the effectiveness of wearing masks for different populations, such as patients with certain underlying conditions, or in different settings, such as on flights and subway. For the current global outbreak of COVID-19, we still recommend that the population should follow the current WHO recommendation or local guidelines. If masks are to be used, they should be combined with hand hygiene and other NPIs to prevent human-to-human mutual infection, and education of wearing masks properly and adherence to other NPIs would be needed. Further large pragmatic trials are also needed to evaluate the efficacy of face mask in preventing respiratory virus transmission in the general population.

## Data Availability

The data used to support the findings of this study are available from the corresponding author upon request.

